# Nursing Documentation Practice and Associated Factors Among Nurses Working in Felege Hiwot Comprehensive Specialized Hospital, Bahirdar Town, North West, Ethiopia, 2025

**DOI:** 10.64898/2026.03.18.26348682

**Authors:** Addisu Endale, Amanuel Taye, Afework Chekol, Alebachew Abat, Sewagegn Zewudie

## Abstract

**Background:** Nursing documentation is an essential component of nursing practice that has a potential to improve patient care outcome. Poor documentation of nursing care activities among nurses has been shown to have negative impacts on the health care quality. The aim of this study was to assess documentation practice and associated factors among nurses working in Felege Hiwot comprehensive specialized hospital from August 1 to August 30, 2025.

**Method:** Institutional based cross sectional study design was employed. The data was checked for completeness, coded and entered in epi –data version 3.1 and analysis was made by STATA version 14. Binary logistic regression analysis was computed to assess associations of factors with documentation practice. Variables with p-value less than 0.25 in Bivariable analysis was entered to final model and P < 0.05 at 95% confidence interval was considered as statistically significant. Odds ratio was used to show strength of association.

**Result:** Out of the 349 respondents, 209 (59.9%) were females. In this study 40.1% of nurses had good documentation practice. Educational level, MSc (AOR, 95%CI; 10.3(3.4-31.8)), attitude (AOR, 95%CI; 2.6(1.5-4.7)), number of patient care (AOR, 95%CI; 5.6(1.9-16.3)) and Knowledge (AOR< 95%CI; 3.7(2.1-6.2)) were statistically associated with documentation practice.

**Conclusion and recommendation:** Poor documentation practice was due to the identified factors. So, it is better to put further effort toward improving documentation practice through providing training on standards of documentation and enhancing the favorable attitude of nurses toward documentation practice.

## Background

Nursing documentation is any written or electronically generated information about a client that describes the care or service provided to that client including what happened and when it happened(1) . It is a main component of safe, ethical and effective nursing practice whether done manually or electronically(2).

Nursing documentation should complete the legal requirements since its consequence may end with malpractice suits(3). A written document is the only evidence if a health professional commits homicide due to negligence. There are also evidences indicating that nursing documentation has relationship with patient mortality (4).

Documentation improve communication between health care provider to facilitate continuity of care and to improve their relationship with patients (5).Additionally, it gives as evidence in legal proceedings through demonstration of the applied nursing knowledge, skills and judgment (6).

Documentation also provides important data for research in nursing in order to improve health outcomes (7). The level of contributions nurses do in the health care system can be witnessed through proper documentation of their roles (8).

A nursing document should be client focused, consists of relevant information, accurate without missing details, chronologically written, clear and concise, permanent, confidential and timely (9).

In Ethiopia, nursing documentation is limited to hand written and are limited critical reflections on the nature and outcomes of nursing care for the patients (8). Even though the quality and effectiveness of nursing practice is mostly demonstrated by documenting the application of the nursing process, nurses may document patient visit registry in the outpatient departments (10).

Documentation is part of their professional obligation, many studies identified deficiencies in practice of documentation among nurses globally. It has been reported that nursing records are often incomplete, lacked accuracy and had poor quality (5, 11).

One of the main problems facing in nursing record is lack of standardization of the documents and forms nurses are required to use. It is not unusual for differently designed forms that have similar functions to exist even within the same facility. Lack of uniformity creates confusion and increases chance of documentation error (12).

The problems for documentation reported so far include inadequate staff, inadequate knowledge concerning the importance of documentation, patient load, lack of in-service training and lack of support from nursing leadership(13). Despite the reasons for not documenting completely and accurately, nurses need to realize that rules and regulations by accrediting organizations expect complete and accurate documentation as indicated in their practice standards (14).

Poor documentation practice in nurses has been shown to have negative impacts on the health care of patients. The impact may lead to harmful consequences like exposing the care provider for medication administration error (2, 12).

In sub Saharan Africa identified deficiencies in various aspects of nursing documentation reported in attitudes, knowledge and practice towards the practice of nursing documentation among nurses (15). Good documentation allows real representation of what is happening on the ground (16).

In Ethiopia, inadequacy of data collection with lack of quality was found to be a problem (17).

## Methods and Materials

### Study design and setting

Institutional based cross-sectional study design was employed .This study was conducted in Felege-Hiwot Comprehensive Specialized Hospital (FHCSH) from August 1 to August 30, 2025. FHCSH is located in Bahir Dar (Capital City of the Region), 565 km away from Addis Ababa in the northwest direction. FHRH is one of the oldest public Hospitals in the country it was established 60 years back and currently providing comprehensive and specialty services, for more than 10 million people of the region.it has more than **865** staffs, among these there are **460** nurses.

### Population

The source population was all staff nurses working in Felege Hiwot Comprehensive specialized hospital. Study population was randomly selected staff nurses working in Felege Hiwot Comprehensive specialized hospital.

### Eligibility criteria

Nurses in-patient (IPD) and out-patient departments (OPD) working in Felege Hiwot Comprehensive specialized hospital were included. Nurses those who are on sick leave, annual leave during the data collection period.

### Sample size determination

The sample size was determine using single population proportion formula by considering; Z=standard normal distribution **(Z=1.96**) taking the **proportion** of good documentation practice as 30.4% from previous study conducted in Amhara region Public Hospital (18), **95%** confidence interval (**CI**), and **5%** margin of error. Thus, the sample size is calculated as follows:

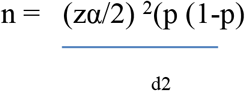

Where n= minimum sample size required for the study P=prevalence/ population proportion **(p=0.304**); d=is a tolerable margin of error **(d=0.05)** = 1.96(1.96) (0.304(1-0.304))/0.05(0.05) = 317.65 => =318 by adding **10%** non-response rate the final sample size is **349**.

### Sampling technique

Simple random Sampling technique was used. List of all nurses was obtained from the nursing leaders (head nurses) of respective wards and the number of samples in each ward was selected according to proportional allocation formula.

**Figure 1.**
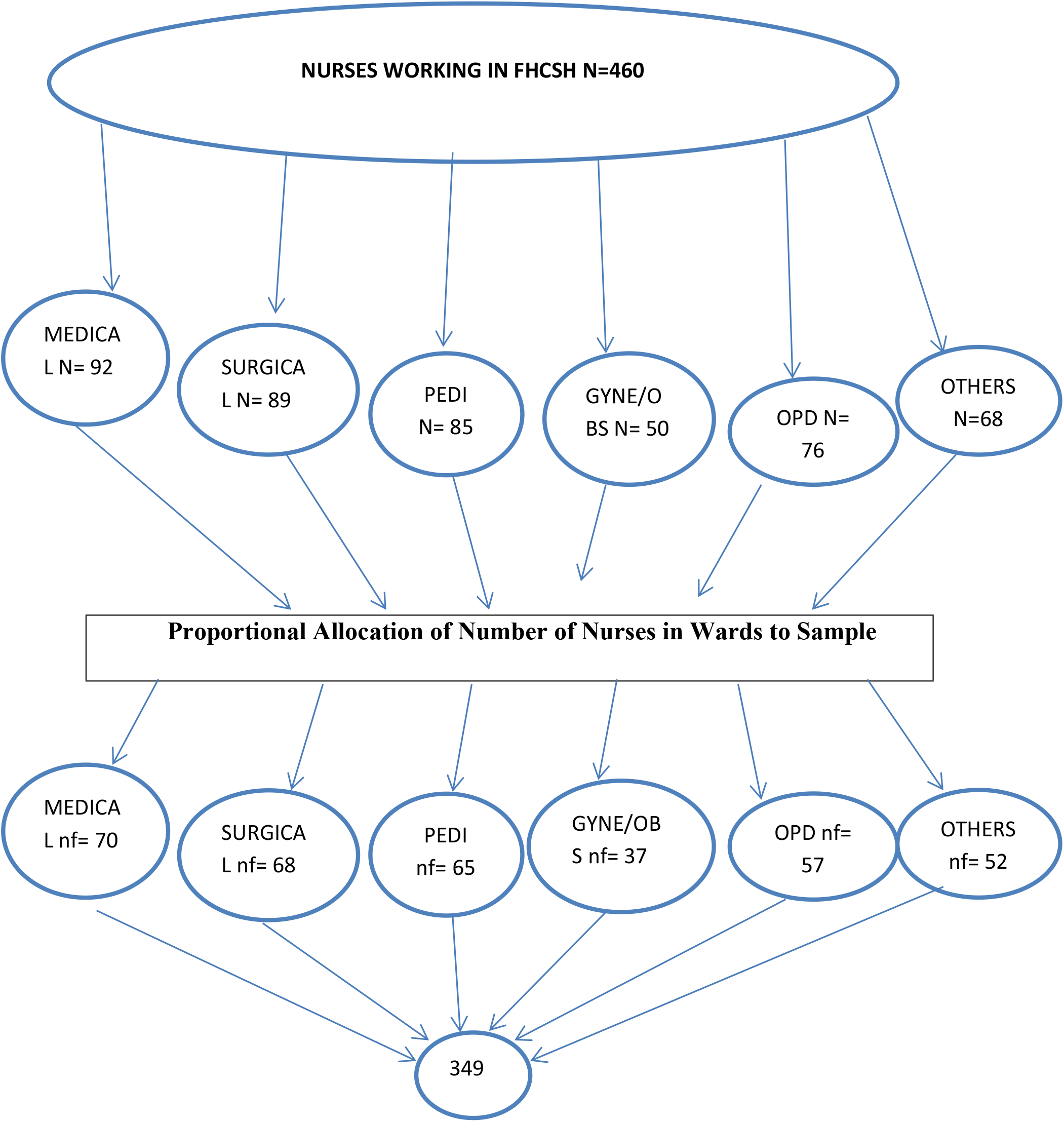
Schematic presentation of sampling procedure for assessment of nursing documentation practice and associated factors among nurses working in FHCSH, Bahir Dar town North West Ethiopia, 2025.

### Variables

Dependent Variable were nurses documentation practice (good/poor) and independent Variables were socio demographic characteristics (age, sex, religion, marital status, income, year of service and level of education, work setting), knowledge towards nursing care documentation, attitude towards nursing care documentation and organizational related factors (inadequate staff, availability of documenting materials, familiarity with documentation guideline, availability of obligation from hospital and motivation from the supervisors, in-service training).

### Operational Definitions

**Good practice**: Those respondents who scored above or equal to the mean score of practice.

**Poor practice**: Those respondents who scored below the mean score of practice.

**Positive attitude:** Those respondents who scored above or equal to the mean score attitude.

**Negative attitude:** Those respondents who scored below the mean score attitude.

**Good knowledge:** Those respondents who scored above or equal to the mean score of knowledge.

**Poor knowledge:** Those respondents who scored below the mean score of knowledge.

#### Data collection procedure and tools

Data was collected by self -administered questionnaire. The data collection tools was in English version. The questions are developed based on literatures related to the topic. Six BSc nurses and one (MSc) supervisors) was recruited for the data collection process and the collector was train for 2 days.

#### Data quality control

Training was provided for data collector prior to the data collection and the collected Data was regularly checked by investigators for completeness and consistency. Its completeness, accuracy and consistency was check prior to entry.

#### Data processing and analysis

The collected data was code and check for completeness. Once data code and check for completeness, data processing will be done. The collected data was checked for completeness and cleanness and entered into Epidata 3.1 then exported to Stata software for analysis. Descriptive statistics was used to organize and summarize the variables. Bivariate analysis for each independent variable with the outcome variable was performed to select candidates for multi variate logistic regression analysis. All independent variables with p-value less than 0.25 was taken as candidates for multivariable logistic regression model then finally p-value of less than 0.05 at 95% CI was used to declare statistical significance. The AOR from multivariate logistic regression was used to measure the strength of association between dependent and independent variables.

## Result

### Socio-demographic characteristics of respondents

Out of the 349 respondents, all returned the questionnaire and the response rate were 100%. From nurses who participated in this study, 209 (59.9%) were females and 105(30.1%) fall within the ranges of 26-30 years age group. Half of the respondents were married. Majority of the respondents 314(90%) orthodox religion followers and more than half of the respondents were holding bachelor degree 200 (57.3%). Almost 140(40.1 %) of the study participants worked as a nurse for 5 years or less (See below).

**Table 1:** Socio demographic characteristics of nurses working in Felege Hiwot Comprehensive Specialized Hospital, Amhara, Ethiopia, 2025 (n=349)

| Variables |  | N | % |
| --- | --- | --- | --- |
| Age group (in a years) | 21 - 25 | 70 | 20.1 |
|  | 26 - 30 | 105 | 30.1 |
|  | 31 - 35 | 105 | 30.1 |
|  | ≥ 35 | 69 | 19.7 |
| sex of respondents | Male | 140 | 40.1 |
|  | Female | 209 | 59.9 |
| Educational level of respondents | Diploma | 63 | 18.1 |
|  | Bachelor degree | 200 | 57.3 |
|  | MSc | 86 | 24.6 |
| Religion of respondent | Orthodox | 314 | 90 |
|  | Muslim | 19 | 5.4 |
|  | Protestant | 16 | 4.6 |
| Marital status | Single | 140 | 40.1 |
|  | Married | 175 | 50.1 |
|  | Divorced | 34 | 9.8 |
| work experience(in years) | ≤5 | 140 | 40.1 |
|  | 6 -10 | 105 | 30.1 |
|  | >10 | 104 | 29.8 |

### Documentation Practice

The mean score for documentation practice were 21 (S.D ± 3.2). Based on this cut-off point, 140 (40.1%) of the study participants had good nursing care documentation practice. (See below)

**Table 2:** Shows documentation practice among nurses working in Felege Hiwot Comprehensive Specialized Hospital, Amhara, Ethiopia, 2025 (n=349)

| **Variables** | **Never** | **Sometimes** | **Always** |
| --- | --- | --- | --- |
| Do you document the assessments you have done for every patient? | 0 | 175 | 174 |
| Do you document problems you find (the nursing diagnosis) for every patient? | 0 | 105 | 244 |
| Do you document the intervention you have done for every patient? | 0 | 105 | 244 |
| Do you document the response to your intervention for every patient? | 34 | 175 | 140 |
| Do you document the fluid you administered to every patient? | 0 | 70 | 279 |
| Do you document the medication you administered to every patient? | 0 | 70 | 279 |
| Do you document the education or advice you have provided to a patient? | 174 | 105 | 70 |
| Do you document the fluid balance status of the patient? | 70 | 209 | 70 |
| Is all your documentation done immediately after care provided to the patient? | 35 | 244 | 70 |

### Knowledge of respondents towards nursing documentation

The mean score for knowledge questions which was 6.6 (S.D ± 2.6). Based on this cut-off point, 175 (50.1%) of the study participants had a good knowledge of nursing care documentation.

Out of total respondents who have participated in the study, majority of them 314 (90%) knew that documentation is a professional responsibility and 244(69.9%) knew that the document should be error free and complete (See table below).

**Table 3:**
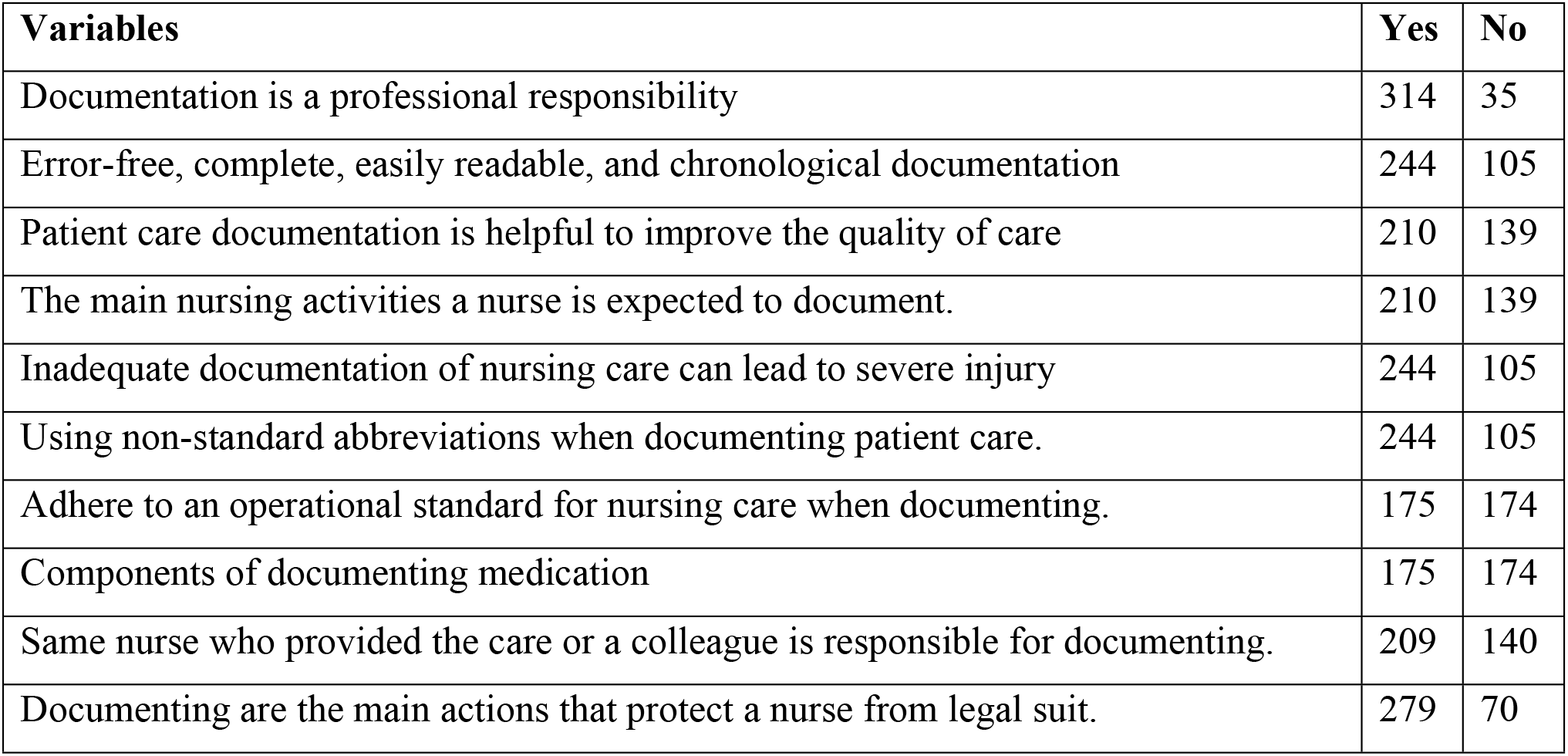
Shows knowledge towards documentation practice among nurses working in Felege Hiwot Comprehensive Specialized Hospital, Amhara, Ethiopia, 2025 (n=349)

### Attitude of nurses towards nursing documentation

Attitudes were assessed via a Likert scale, with scores ranging from strongly agree (5) to strongly disagree (1).The total mean score were 33 (S.D ± 8.9). Among all respondents, one third 105(30.1%) agreed documentation value for the hospital quality; half of the respondents were neutral for documentation has a positive impact on patient safety; 104(29.8%) were strongly disagree written can replace oral shift report. All over were assessed and indicated that 70.2%(245) had positive attitude (see below).

**Table 4:**
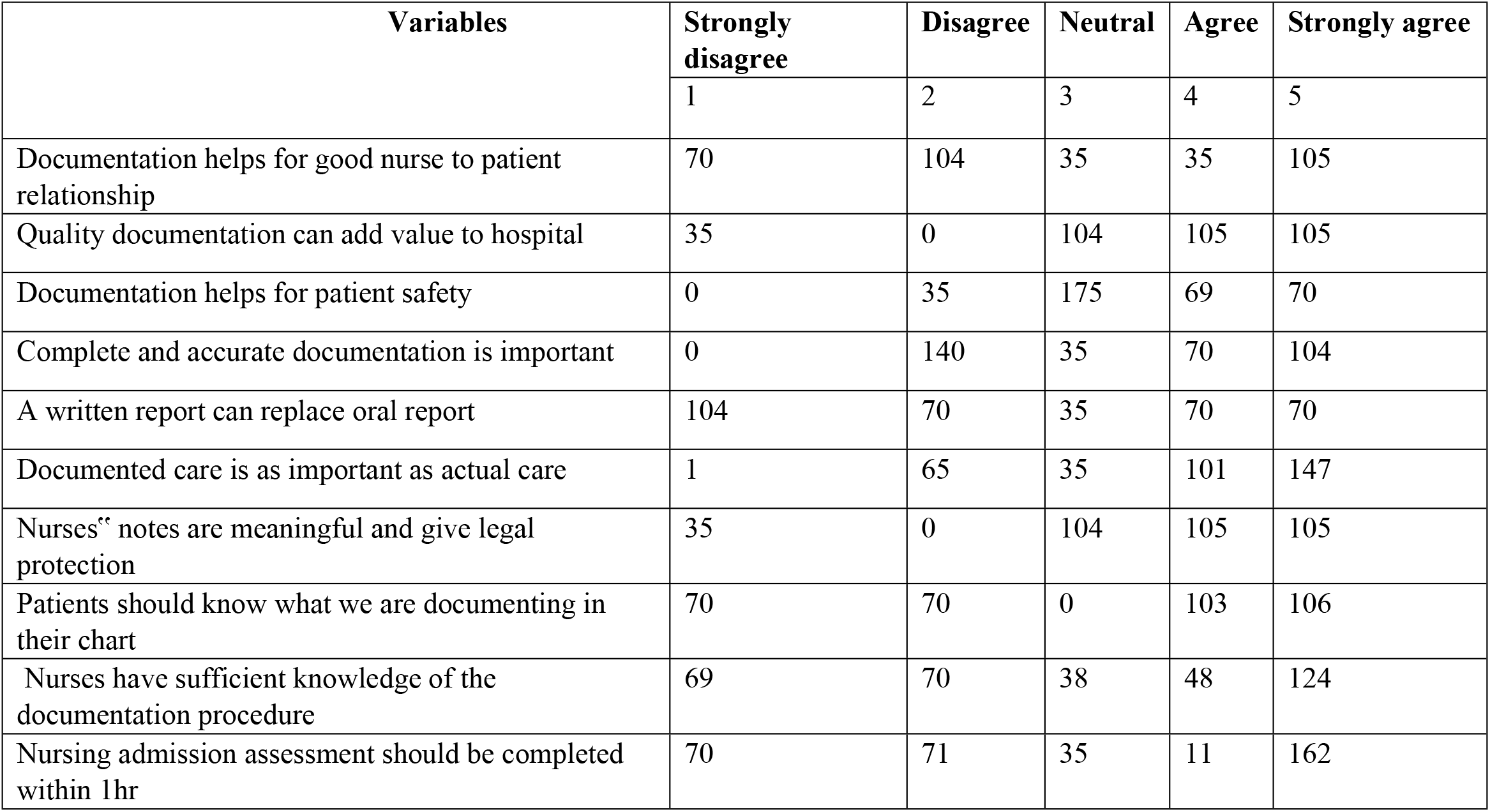
Shows attitude towards documentation practice among nurses working in Felege Hiwot Comprehensive Specialized Hospital, Amhara, Ethiopia, 2025 (n=349)

### Organizational related description

Among study participants, majority 279(79.9%) were participated in service training. More than half, 209(59.8%) had no access documentation guideline and also 279(79.9%) had no adequate time for documentation (see below).

**Table 5:** Shows organizational related description among nurses working in Felege Hiwot Comprehensive Specialized Hospital, Amhara, Ethiopia, 2025.

| Variables |  | Frequency | Percent |
| --- | --- | --- | --- |
| Are you attended in-service training about documentation? | Yes | 279 | 79.9 |
|  | No | 70 | 20.1 |
| Average number of patients cared for by a nurse per shift | <=5 | 104 | 29.8 |
|  | 5 - 10 | 245 | 70.2 |
| Is their documentation guideline availability? | Available | 140 | 40.1 |
|  | Not available | 209 | 59.9 |
| Are you Familiar with documentation standards? | Yes | 209 | 59.9 |
|  | No | 140 | 40.1 |
| Is Availability of documentation sheet adequate? | Yes | 139 | 39.8 |
|  | No | 210 | 60.2 |
| Is their Motivation from supervisor? | Yes | 34 | 9.8 |
|  | No | 315 | 90.2 |
| Is time adequate? | Yes | 70 | 20.1 |
|  | No | 279 | 79.9 |

### Factors associated with documentation practice

For analysis of the data, bivariate and multi variate logistic regression were done by using binary logistic regression. Crude and Adjusted odds ratio with 95% confidence interval was calculated to determine the strength of association and statistical significance between documentation practice and each independent variable. The analysis described that educational level, attitude, knowledge and number of patient care strongly associated with poor documentation practice and statistically significant.

**Table 6:** Shows Bi variable and multivariable logistic regression analysis of factors associated with documentation practice among nurses working in Felege Hiwot Comprehensive Specialized Hospital, Amhara, Ethiopia, 2025.

| Variables | Documentation practice |  |  |  |  |
| --- | --- | --- | --- | --- | --- |
|  | Good<br>(N=140) | Poor<br>(N=209) | COR (95%CI) | AOR (95% CI) | P-value |
| **Educational level** |  |  |  |  |  |
| Diploma | 26 | 37 | 1.8(1.0-3.2) | 2.2(1.1-4.5) | 0.03** |
| Bachelor | 57 | 143 | 1 | 1 |  |
| MSC | 57 | 29 | 4.9(2.9-8.5) | 10.3(3.4-31.8) | 0.00** |
| **Number of patient care** |  |  |  |  |  |
| ≤5 | 35 | 69 | 1 | 1 |  |
| 6 – 10 | 105 | 140 | 1.5(0.9-2.4) | 5.6(1.9-16.3) | 0.00** |
| **Attitude** |  |  |  |  |  |
| Negative | 70 | 175 | 0.2(0.1-0.3) | 2.6(1.5-4.7) | 0.00** |
| Positive | 70 | 34 | 1 | 1 |  |
| **Knowledge** |  |  |  |  |  |
| Poor | 35 | 139 | 0.2(0.1-0.3) | 3.7(2.1-6.2) | 0.00** |
| Good | 105 | 70 | 1 | 1 |  |
| **Time adequacy** |  |  |  |  |  |
| Yes | 35 | 35 | 1 | 1 |  |
| No | 105 | 174 | 1.7(1.0-2.8) | 0.6(0.3-1.4) | 0.21 |

## Discussion

Poor documentation in nurses has been shown to have negative impacts on the health care of patients, the health care providers and on the profession in general. Various studies have shown that documentation problem is still a critical issue in both developed and under developed countries, especially in Sub-Saharan Africa including Ethiopia. In this study, 140(40.1%) had good documentation practice (95% CI; 34.9-45.3). It is nearly similar with the study conducted in Indonesia, but lower than finding in Nigeria(19, 20). Similar finding in Ethiopia slightly higher than the current finding (18). This discrepancy might be due to staff educational level difference, accessibility of training and workload variation.

This study showed that nurses those had master 10 times more practice documentation than bachelor. But, those had diploma nearly 2 times poor documentation practice. This might showed that upgrading their level improve documentation practice.

According to this finding, those nurse had more number of patient care had 5.6 times poor documentation than those who had low in number. This is similar with the study conducted in Iran and Nigeria (20, 21). This might indicated that workload hinder documentation practice and taking their time for saving of patients.

This finding indicated that nurses had negative attitude had 2.6 times more poor documentation practice. This is similar with study conducted in Uganda and Gondar, Ethiopia (22, 23). This might due to those nurses had positive attitude thought it will beneficiate both client and health institution.

This study reported that nurses had poor knowledge 3.7 times more poor documentation practice. This is similar with finding in Netherland, Ghana, Uganda and Ethiopia (18, 21, 24, 25). It might indicate being knowledgeable on documentation favor their practice.

## Limitation of the study

- The responses might have been liable social desirability bias.
- Self-report may over/underestimation the level of documentation practice

## Conclusion and recommendation

Nursing documentation practice was poor among nurses under the study. Educational level, attitude, knowledge and number of patient care were significantly associated with practice of nursing care documentation.

It has been accepted that nursing documentation is a very important aspect of professional practice to nurses. Based on the finding of this study, the following recommendations are forwarded for:

❖ **For staff nurses:** To improve their documentation practice by upgrading their educational level, improve their knowledge and attitude.
❖ **For Hospital:** Provide sustained continuing long term training and reduce workload of nurse by maintaining adequate manpower.
❖ **For researchers:** Considering longitudinal study to examine actual practice and to carry out large scale studies in order to address the problem in wider context.

## Data Availability

All relevant data are within the manuscript and its Supporting Information files.

## Abbreviations and acronyms

ANA: American Nurses Association
AOR: Adjusted Odds Ratio
COR: Crude Odds Ratio
EHRIG: Ethiopian hospital reform implementation guideline
FDRE: Federal Democratic Republic of Ethiopia
FHCSH: Felege Hiwot Comprehensive Specialized Hospital
GYN/OBS: obstetrics and gynecology
HMIS: Health Management Information System
MSC: Master of Science
OPD: Out Patient Department
PED: Pediatrics
SPSS: Statistical Package for Social Sciences
WHO: World Health Organization

## Acknowledgements

We would like to thanks data collectors who participated in the study.

## Authors’ contributions

AE conceptualization, methodology, software analysis, drafting and edition of manuscript. AT, AF, AA and SZ supervision, validation, reviewing and editing.

## Availability of data and material

The data set are available in the corresponding author on reasonable request.

## Ethical consideration

Ethical clearance was obtained from Amhara public health institute. A formal written letter was provided to specialized hospitals. To kept confidentiality the name of nurses was not recorded and their information was not disclose to third part.

## Consent for publication

Not applicable

